# Cracks in the Foundation: How Data-Hungry and Sensitive to Domain Shift are Vision Foundation Models for Computational Pathology?

**DOI:** 10.64898/2026.01.06.25342815

**Authors:** Anja Witte, Patrick Fuhlert, Maximilian Lennartz, Jan Baumbach, Tobias B. Huber, Ewert Bengtsson, Guido Sauter, Marina Zimmermann, Stefan Bonn

**Author notes:** Corresponding author.:Email address (Stefan Bonn). These authors contributed equally to this work (Anja Witte, Patrick Fuhlert). These authors contributed equally to this work (Marina Zimmermann, Stefan Bonn).

## Abstract

**Background:** Vision Foundation Models (VFM) have emerged as a promising approach for computational pathology, offering scalable feature representations that may reduce labelled-data requirements and improve robustness to variation in tissue preparation and digitisation. However, VFM decoder and dataset size requirements as well as the performance under real-world domain shifts remain unclear.

**Methods:** We evaluated six contemporary VFMs on a protocol-variant Prostate Cancer (PCa) dataset comprising 37 683 tissue microarray spot images from 10 412 patients. The dataset includes six controlled domain shifts arising from differences in staining duration, section thickness, scanner type, and sampling location. Two clinically relevant downstream tasks were examined: ISUP grading and 5-year relapse prediction. We compared two decoder architectures, quantified dataset-size requirements using a saturation analysis (45–5727 samples), and assessed cross-domain robustness using out-of-domain test sets.

**Findings:** Larger VFMs consistently outperformed smaller models in peak accuracy and robustness metrics. Contrary to expectations of data efficiency, all models showed strong dependence on training-set size, requiring at least 1000 samples to approach stable results. All VFMs showed notable degradation under protocol-level domain shifts, with performance reductions of 4 to 13 percentage points in both cancer grading and relapse prediction, although larger models exhibited somewhat greater robustness. Furthermore, KNN-based probing performed substantially worse than a decoder-based approach across all architectures.

**Interpretations:** Despite their strong representational capacity, current VFMs do not yet provide reliable domain generalisation or data-efficient performance in computational pathology. Decoder design remains essential, and substantial amounts of labelled data are still required to achieve clinically meaningful accuracy. Further advances in pre-training strategies, decoder architectures, and domain adaptation methods will be crucial for translating VFMs into robust clinical tools.

**Research in context:** *Evidence before this study:* This study focuses on pathology foundation models, which offer promising improvements in performance, data requirements, and robustness to domain shifts for computational pathology. To identify relevant studies, we searched in Google Scholar for research published before 1 April 2025. We searched for studies introducing novel foundation models trained on pathology images, or reviews comparing those models in terms of performance or robustness. The search terms used were ‘computational pathology’, ‘benchmarking’, ‘review’ and ‘foundation model’, as well combinations of these terms. We found that many studies focus on increasing the complexity of pathology foundation models while using increasingly extensive and heterogeneous pre-training datasets. Various benchmarking studies demonstrate the superior performance and robustness of more recent and larger foundation models. However, these studies have limitations in their evaluation datasets. Either they cover only a domain shift due to a different scanner device, or they have small sample sizes. We also identified a research gap regarding the requirement for large datasets to train a decoder based on a pathology foundation model for a specific downstream task.

*Added value of this study:* The goal of this study was to evaluate the necessity of large downstream task datasets and the domain shift robustness of multiple pathology foundation models. For this purpose, we used our internal protocol-variant prostate cancer dataset, which provides a controlled evaluation setup as multiple domain shift types have been intentionally and separately introduced for different sub-datasets. Our saturation analysis revealed that at least 1000 samples were necessary to achieve good performance. Furthermore, our findings show that none of the evaluated foundation models are robust against all of our domain shifts, though larger models generally perform better.

*Implications of all the available evidence:* This study reveals that increasing the capacity of pathology foundation models improves performance and robustness. However, we demonstrated that all models exhibit some degree of performance degradation for certain domain shifts and require substantial datasets for training on downstream tasks. These limitations demonstrate that pathology foundation models do not fully address the issues of robustness and data requirements.

## 1. Introduction

Computational pathology is undergoing a rapid transformation driven by advances in Artificial Intelligence (AI) with VFMs emerging as a promising paradigm for histopathological image analysis. By pre-training large Vision Transformer (ViT) architectures on vast collections of Whole Slide Images (WSIs), VFMs aim to provide generalizable, biologically meaningful feature representations that can be adapted to diverse downstream tasks with minimal labelled data. These approaches have demonstrated impressive performance across cancer classification, grading, and prognostication tasks, and are increasingly proposed as a route to more robust and scalable computational pathology systems.^1, 2, 3^

Despite this progress, the reliability of VFMs in real-world clinical environments remains uncertain. Digital pathology workflows are characterised by substantial variability in tissue processing, staining protocols, section thickness, and scanner hardware.^4^ These technical differences introduce domain shifts that can alter colour, texture, and structural appearance in ways unrelated to underlying biology, posing a major challenge for generalization. Although modern VFMs often claim improved robustness based on diverse pre-training datasets and self-supervised learning, the extent to which these benefits translate into practical resilience against protocol-level variation has not been comprehensively evaluated. Existing benchmarking efforts typically compare models across institutions without knowledge of the underlying acquisition differences,^5, 6^ or rely on small scanner-shift datasets that capture only a narrow spectrum of real-world variability.^7, 8^

Another open question concerns data efficiency. VFMs are frequently promoted as requiring “little data” for fine-tuning,^1^ allowing smaller institutions or rare-disease cohorts to benefit from high-performance AI tools. However, downstream performance does not only depend on the encoder, but also on decoder architecture, training strategy, and inherent task difficulty. What constitutes “little data” in computational pathology remains unclear, and systematic investigations into dataset-size requirements for VFM-based workflows are scarce.

To answer these pertinent questions, we conducted a comprehensive evaluation of six contemporary VFMs using a large, protocol-variant PCa dataset intentionally designed to capture real-world clinical data variability. The dataset includes 69 251 tissue microarray images from 17 700 patients, spanning six controlled domain shifts, each reflecting a single variation in staining duration, tissue thickness, scanner type, or anatomical sampling. This setting allows rigorous examination of model robustness while controlling for patient-level biological factors. Furthermore, we systematically analysed decoder requirements and dataset-size dependencies through a saturation study ranging from 45 to 5727 training samples. VFM performance was assessed on two clinically relevant tasks: ISUP^9^ cancer grading and 5-year relapse prediction.^10, 11^

Our study provides three key insights. First, although VFMs have strong representational capacity, they typically depend on specialized decoder architectures to achieve high-quality downstream predictions. Second, all models, even billion-parameter transformers, showed significant data hunger for training the decoder architecture. Strong performance requires roughly 1000 samples or more. Third, none of the models displayed consistent robustness to protocol-level domain shifts, with performance dropping markedly on out-of-domain subsets. Together, these findings challenge common assumptions about the robustness and data efficiency of current VFMs and highlight the need for next-generation training paradigms, decoder designs, and domain generalization strategies before such models can serve as dependable tools in computational pathology.

## 2. Methods

### 2.1. Vision Foundation Model Selection

The main differences between the selected VFMs are model architectures, and pre-training datasets. Earlier VFM publications like `CTransPath`^12^ and `REMEDIS`^13^ proposed Convolutional Neural Network (CNN)-based architectures. Following the emergence of ViTs and their superior performance compared to CNNs, recent research has utilised encoders based on these architectures.^1^ The commonly used ViT-based encoder models range from ViT-Small with 22M parameters to ViT-Giant with 1·1B parameters. To accommodate the variation in model complexity, different pretraining datasets are employed across the approaches. The number of training WSIs commonly ranges from thousands to millions, with larger datasets typically utilized for pre-training more extensive encoders. To enable VFMs to learn general pathology domain knowledge, self-supervised learning methods are commonly used. Among these, `DINOv2`^14^ is the most frequently adopted approach to date. As shown in table A.3, numerous VFMs for pathology have been published in the latest years, whereas a trend towards larger models is visible. In order to select VFMs that are comparable and represent the current research, we defined the following selection criteria:

1. A main aim of this work is to investigate the impact of model size on robustness and at least one model of each category ViT is chosen.
2. The selected model must be available on Hugging Face.^3^
3. To enhance comparability, models trained with similar pre-training algorithm and datasets are preferred.
4. In case of multiple versions of a model, the most recent one (as of 1. April 2025) is selected.

Following the pre-defined selection criteria, we decided on the VFMs presented in table 1. As Kaiko models^15^ are available in multiple sizes and pre-trained similarly, they are chosen for ViT-Small, ViT-Base and ViT-Large. For ViT-Huge, two models were selected. UNI2^1^ has shown good performance in other benchmarks. Further, Virchow-2^2^ represents the model with the largest pre-training dataset (3·1M WSIs) and is the only model that suggests combining the patch-level tokens with the class token. Lastly, H-optimus-1^16^ was chosen as it is the largest model available on https://huggingface.co/ at that time.

**Table 1:** An overview of the compared VFMs. Architectures with ^∗^ have modified numbers of layers or attention heads. *#WSIs* specifies the number of images that have been used to pre-train a VFM. The column *Dim* specifies the output embedding dimension for each VFM. Unless the VFM authors suggest otherwise, this represents the Class Token (CLS) embedding. VFMs marked with ^+^ concatenate the CLS token and the patch embeddings. The columns *#Encoder* and *#Decoder* represent the corresponding number of parameters for each VFM and its decoder, respectively. These numbers are presented separately because the encoder is frozen during our experiments and only the decoder is trained. *#Total* shows the sum of *#Encoder* and *#Decoder*, representing the total number of parameters for an architecture.

| Model Name | Year | Architecture | Method | #WSIs | Dim | #Encoder | #Decoder | #Total |
| --- | --- | --- | --- | --- | --- | --- | --- | --- |
| Kaiko-S<sup>15</sup> | 2024 | ViT- Small/16 | DINO | 29k | 384 | 22M | 68.1k | 22M |
| Kaiko-B<sup>15</sup> | 2024 | ViT- Base/16 | DINO | 29k | 768 | 86M | 172k | 86M |
| Kaiko-L<sup>15</sup> | 2024 | ViT- Large/14 | DINOv2 | 29k | 1024 | 304M | 262k | 304M |
| Virchow2<sup>2</sup> | 2024 | ViT- Huge/14 | DINOv2 | 3.100k | <sup>+</sup>2560 | 632M | 1.100k | 632M |
| UNI2-h<sup>1</sup> | 2025 | *ViT- Huge/14 | DINOv2 | 350k | 1536 | 681M | 492k | 681M |
| H-optimus-1<sup>16</sup> | 2025 | ViT- Giant/14 | DINOv2 | 1.000k | 1536 | 1.100M | 492k | 1.100M |

### 2.2. Dataset

The internal dataset contains 69 251 Tissue Microarray (TMA) spot images provided by the Pathology Institute of the University Medical Center Hamburg-Eppendorf (UKE). The images are collected from 17 700 patients that underwent Radical Prostatectomy (RP) between 1992 and 2014. Tissue preparation and digitization process follows six different protocols to emulate different domain shifts resulting from different clinics. Accordingly, our dataset comprises six different sub-datasets of which we show samples in the data section of fig. 1.

**Figure 1:**
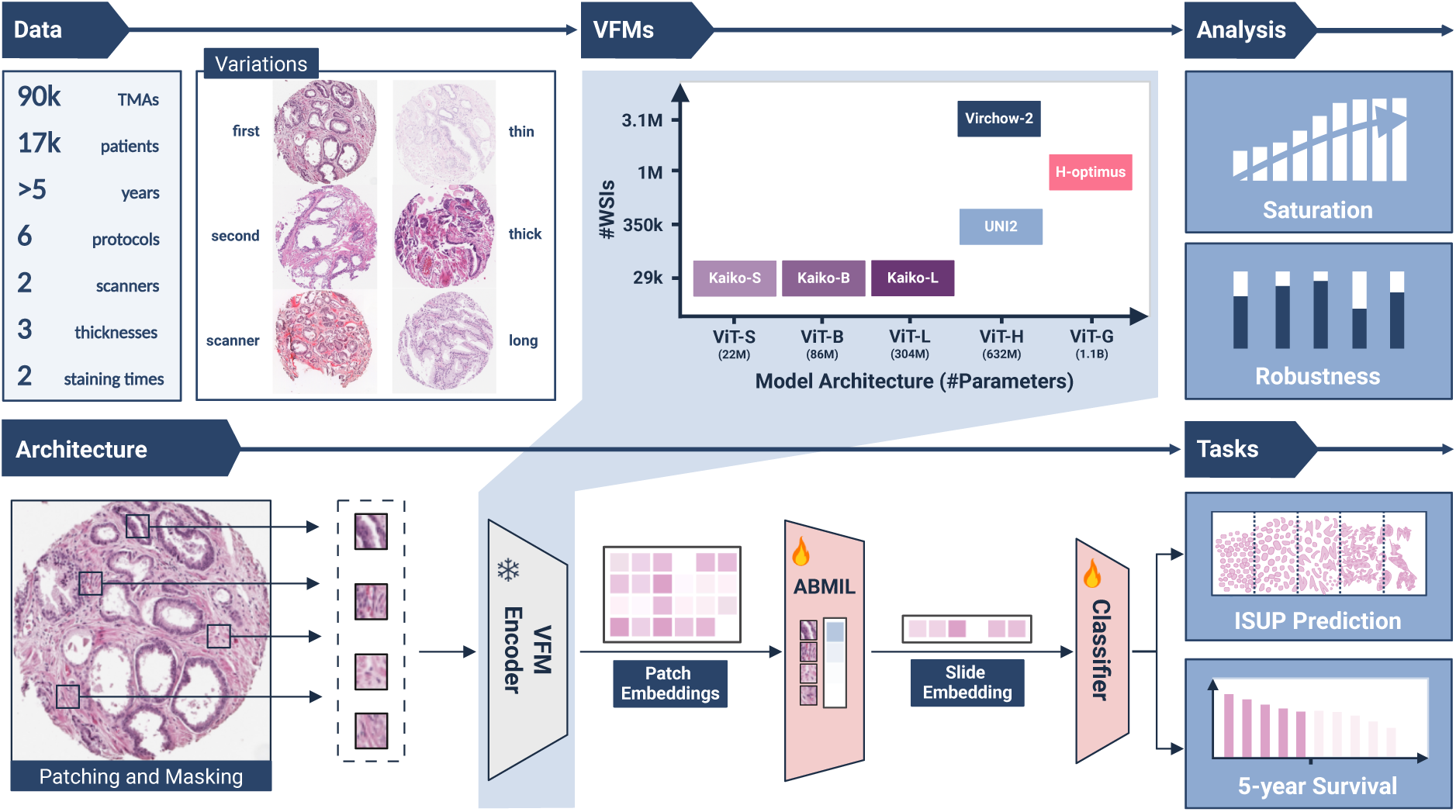
Overview of this work’s benchmarking approach. The frozen parts of the network architecture are depicted in blue, while trained parts are shown in red. Each WSI is divided into non-overlapping tiles. All tiles showing tissue are encoded using six different VFM encoders. The resulting tile embeddings for each VFM are then aggregated into a slide-level representation using ABMIL. Finally, a classifier is trained to predict the outcomes of our two downstream tasks: ISUP classification and 5-year survival prediction. We conduct a saturation analysis with this architecture to determine the required number of samples. Additionally, we assess the robustness of the trained models using our internal dataset.

In this study, we evaluate our methods on two tasks: ISUP grading and 5-year relapse prediction. In addition to the histological images, the dataset includes patient-specific information that we use as ground truth. The included ISUP gradings are determined based on examinations of the entire prostate by pathologists within the UKE. Moreover, high-quality follow-up data regarding various relapse-related events are available. The events of interest are biochemical recurrence, additional unplanned therapy, metastasis, and PCa-related death. Following recent research, the patient is considered to have experienced a relapse if any of these events occur within five years after RP.^17^

The dataset is filtered accordingly to only include patients with at least five years of documented follow-up or one of the previously mentioned adverse events. Similarly, we exclude patients without ISUP grading and those who received adjuvant treatment. Finally, after removing images with insufficient quality, our final dataset contains a total of 37 683 images from 10 412 patients.

After filtering, the main sub-dataset UKE.first provides 8141 TMA spots following the standard digitization protocol of the UKE. It contains a selected TMA spot that is most representative for the disease progression of the individual patient. Tissue samples were sliced at 2·5 µm, stained with Haematoxylin and Eosin (H&E) for 4:00 and 1:20 minutes respectively, and digitized by an Aperio scanner under a magnification of 40x (0·25 µm/px). Note that the following sub-datasets show negligible biases in terms of patient distribution as shown in table A.2. Additional sub-datasets focus on single variations in the digitization protocol. The UKE.second (*n* = 1164) sub-dataset contains a secondary TMA spot that was obtained from another part of the cancerous area of the prostate for the same patient. UKE.scanner (*n* = 1208) is a collection of spots that were scanned by a different scanner vendor, i.e. 3DHistech at 80x magnification (0·125 µm/px). UKE.thin (*n* = 885) includes thinner cut spots of 1 µm instead of 2·5 µm used in the standard protocol. In contrast, spots of the UKE.thick sub-dataset (*n* = 891) were cut thicker at 10 µm. Lastly, UKE.long (*n* = 893) contains longer stained TMA spots at 40 min haematoxylin and 10 min eosin staining respectively.

For model development, the UKE.first sub-dataset is split into training, validation, and test set stratified at patient level and 5-year survival endpoint to avoid leakage of patient-level information. Further, we construct test sets for the other sub-datasets to include only patients of the UKE.first test split.

### 2.3. Experimental Setup

We consider two types of tasks for prostate cancer risk assessment: ISUP grading and 5-year relapse prediction. ISUP classifies cancerous tissue into one of five grades based on glandular morphology and reflects the histopathological severity of the disease. Patient-level expert annotations within our internal UKE dataset serve as ground-truth. Model performance for this task is quantified using Cohen’s kappa (Cohen’s *κ*), which accounts for chance agreement between predicted and true grades.^18^ Relapse prediction is formulated as a binary classification problem, defined by the occurrence of relapse-related events as described in section 2.2 within five years after RP. Performance is evaluated using Area under the Receiver Operating Characteristic Curve for 5-year relapse prediction (AUROC5), providing a threshold-independent measure of discriminative ability.

All experiments are conducted on the six sub-datasets derived from the main cohort described in section 2.2, namely UKE.first, UKE.second, UKE.scanner, UKE.thin, UKE.thick, and UKE.long. Of these, only samples of UKE.first were used to train the model (on *n* = 5727 TMAs), while the remaining sub-datasets were exclusively employed for testing in both downstream tasks.

### 2.4. Training Regime

All VFMs are fine-tuned and evaluated with the same workflow as shown in fig. 1. The input images are resized to represent a magnification of 40x (0·25 µm/px). Each image is then patched into equally sized and non-overlapping tiles. The tile size is 224 × 224 × 3 pixels. To remove background tiles that show less than 10 % of tissue, filtering based on Otsu’s thresholding^19^ is applied. The resulting tiles with at least 90 % of tissue content are then normalized according to the individual encoders. Each tile is separately encoded for each selected VFM where the embedding dimension *d* depends on the encoder architecture. The extracted tile embedding has a shape of *n* × *d* which is aggregated into a bag-level representation of the overall TMA of shape 1 × *d* using Attention-Based Multiple-Instance Learning (ABMIL).^20^ Finally, this vector is passed to a task-specific classification head of two dense layers with a hidden dimension of 128 nodes to determine the final prediction. While training the architecture on our downstream tasks, the pre-trained feature encoder is frozen and we only train the Multiple Instance Learning (MIL) layer combined with the task-specific head. Depending on the tile-level embedding dimension *d* of the respective foundation model, this leads to a comparatively small number of trainable model parameters ranging from 68k for Kaiko-S to 1·1M for Virchow-2 (see table 1) depending on the embedding dimension *d*.

The individual evaluations undergo training up to 25 times to measure the performance variance. We use five different seeds for weight initialization and, if necessary, five more for training data sub-sampling. A batch size of 128 is used as long as at least two batches remain, otherwise the batch size is halved. Further, `AdamW`^21^ is chosen as the optimizer with a weight decay of 0·01, and early stopping with a patience of 20 epochs for a maximum of 40 epochs is used along with a cosine annealing learning rate scheduler. Each model’s learning rate is tuned separately with Bayesian search over 20 runs (see table A.4).

## 3. Results

In this section, we present a comprehensive evaluation of six foundation models for prostate cancer grading. To this end, a total of 37 683 TMA spot images from 10 412 patients met the eligibility criteria and were included in the evaluation. Model performance was assessed across in-domain data and five protocol-specific sub-datasets representing scanner variation, section thickness, staining duration, and sampling location (details in section 2.2). Performance was evaluated for clinically relevant ISUP grading and 5-year relapse prediction with an emphasis on the impact of architectural choice, training set size, and cross-domain generalization.

### 3.1. Task-specific decoders increase VFM performance

Foundation models are pre-trained on extensive and diverse datasets and considered to provide biologically informative image embeddings that could be suited for prediction and classification tasks without further training. To understand if the six VFMs tested in this study contain meaningful embedding representations, we perform a dense feature similarity analysis for each model (see fig. 2a). By using three patches representing different image structures, namely glandular tissue, stroma and background, we can qualitatively show that all models except Virchow-2 cluster those sample structures within their embedding spaces. UNI2 and H-optimus-1 perform well at separating these structures. In contrast, Virchow-2 focuses on tile-level information. The results indicate that for most VFMs biological information is relevant for the embedding space structure, which is relevant for zero- and few-shot learning.

**Figure 2:**
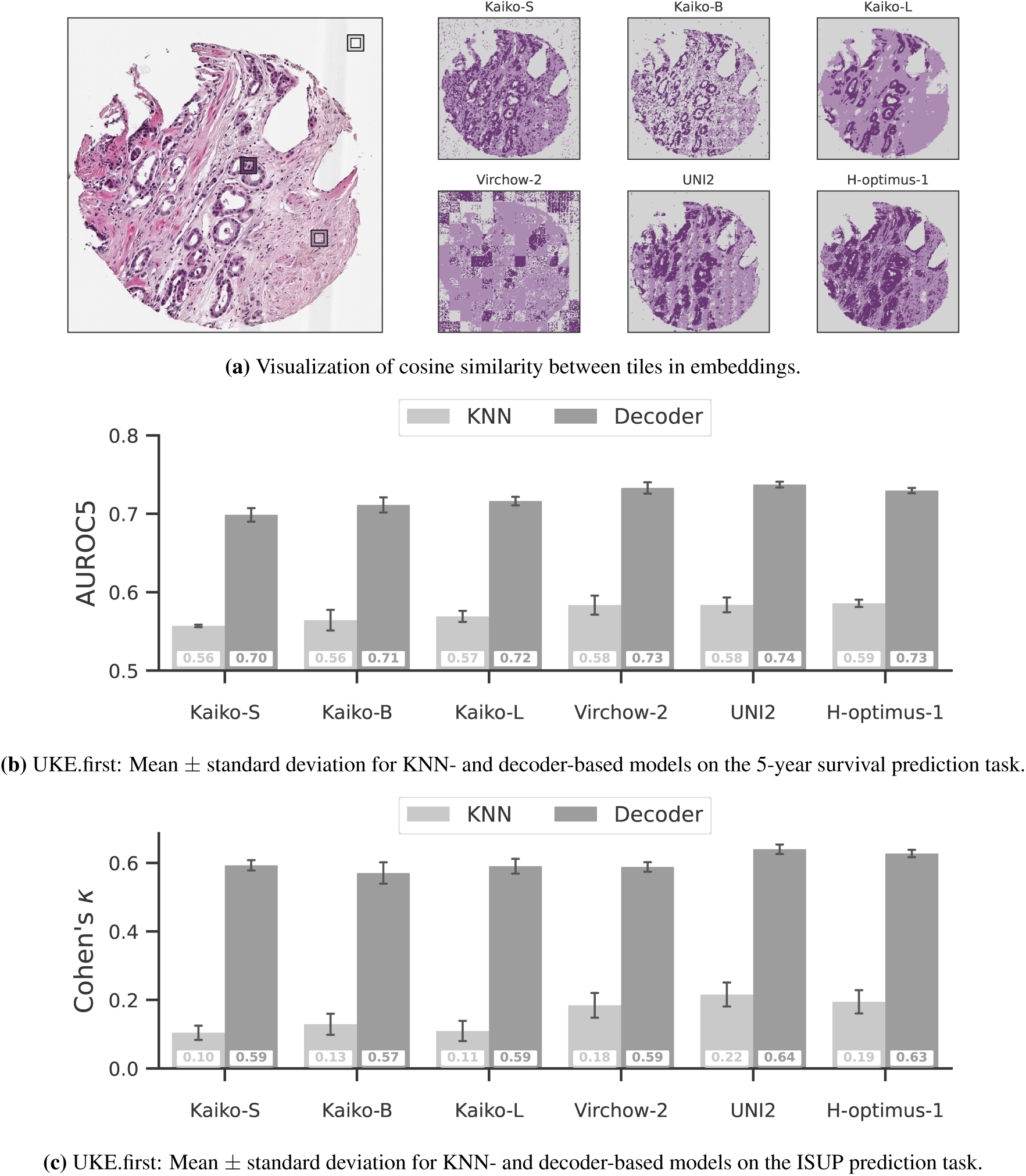
Embedding evaluation. (a) Visualizing of embeddings by probing three reference minipatch embeddings representing glandular tissue (dark purple), stroma (light purple), and background (gray) for the six analysed VFMs. Each minipatch’s colour corresponds to the closest of those three classes based on cosine similarity of the minipatch embedding. Comparing KNN and decoder-based results on UKE.first for 5-year relapse prediction (b), and for ISUP classification (c).

To investigate whether these embedding spaces are advantageous for our downstream tasks, 5-year relapse prediction and ISUP grading, we perform KNN probing as a common technique for evaluating embeddings learned through self-supervised methods.^14, 22, 1^ In KNN probing, a prediction is inferred based on the labels of its *n* closest neighbours. In our case, we use the Euclidean distance to identify *n* = 20 neighbours based on the aggregated bag embedding. This simple method needs no training and completely relies on the encoder embeddings. As previous research has shown, a decoder architecture that is more advanced and has undergone training achieves good performance on 5-year relapse prediction.^11^ We therefore compare KNN probing with our proposed decoder architecture (see section 2.4). This allows us to understand the embedding’s relevance towards our downstream tasks, namely ISUP classification and 5-year relapse prediction.

For 5-year relapse classification (fig. 2b, table B.5a), decoder performance reached AUROC5 values of 0·70 to 0·74 compared to 0·56 to 0·59 for KNN probing, which led to an increase of 14 to 16 percentage points (pp). The smallest model with the least parameters, Kaiko-S, showed the lowest performance (0·56 KNN, 0·70 decoder). The models with the most parameters, UNI2 (0·58 for KNN, 0·74 for the decoder) and H-optimus-1 (0·59 KNN, 0·73 decoder), achieved the best AUROC5 performance for KNN and decoder predictions.

Similarly, for ISUP classification (fig. 2c, table B.5b), all decoder models substantially outperformed KNN probing across all six VFMs with an increase of 41 to 49 pp. For KNN-probing, the worst Cohen’s *κ* was found for Kaiko-S (0·10) while UNI2 (0·22) reached best performance. For our tailored decoder architecture, worst and best Cohen’s *κ* was achieved by Kaiko-B (0·57) and UNI2 (0·64) respectively. Overall, the smaller kaiko models improve by 44 to 49 pp while the larger models improve by 40 to 43 pp in terms of Cohen’s *κ* when replacing KNN-probing with the tailored decoder architecture.

These results for ISUP classification and 5-year relapse prediction indicate that all VFMs require task-specific decoders to achieve good performance where the boost in performance is even bigger for the smaller VFMs. In the following, only results from such a tailored decoder architecture are presented.

### 3.2. Model fine-tuning requires large training data

The previous section highlights that large VFMs provide good data representations, yet task-specific decoder training is required for high fidelity. This raises the question of how much data is required for decoder training to reach good performance, in settings of varying task complexity. To address this question, we conduct a saturation analysis for both 5-year relapse prediction (easier task) and ISUP classification (harder task) on progressively smaller subsets of our training set, which originally consists of 5727 TMAs from UKE.first. The size of the training set was gradually reduced by half, with a minimum of 45 samples after eight iterations. For each step, we sub-sampled the training datasets five times to estimate the performance variance. The validation and test datasets remain the same for all experiments. We consider performance to be quasi optimal once it is within 1 pp of the highest achieved mean AUROC5 (5-year relapse prediction) or 2 pp of the highest Cohen’s *κ* (ISUP prediction). We consider the standard deviation to be quasi optimal when it falls below 0·01 for AUROC5 or 0·02 for Cohen’s *κ*.

The results in fig. 3a (see also table B.6a) show that the mean 5-year relapse prediction performance on UKE.first improves with additional training data for all analysed VFMs, indicating that saturation is not yet achieved for this dataset. For the three smaller Kaiko models, increasing the training dataset from 45 to 5727 samples elevates AUROC5 by 8 pp from 0·63 to 0·64 to 0·71 to 0·72. This performance gain with increasing data size is 4 to 6 pp for the models with more parameters. The performance of Virchow-2, for instance, increases by 6 pp (0·67 to 0·73) in AUROC5 and H-optimus-1 by 4 pp (0·69 to 0·73). UNI2 achieves the best performance for each analyzed training data subset totalling a gain of 5 pp (0·69 to 0·74). Overall, there is a trend that the AUROC5 standard deviation decreases for all models when the number of training samples is increased. The standard deviation for 45 training samples ranges from 0·017 to 0·034 and gets decreased to 0·002 to 0·007 when all 5727 training samples are used (see table B.6a for details). A substantial drop is visible for 1432 training samples where it drops below 0.01 for all models except Kaiko-B (0.011). Overall, all analysed models perform better and become more reliable with more training data, particularly with more than 1000 samples. Comparing the models to each other demonstrates superior performance of the larger models, especially UNI2 that outperforms all other VFMs, whereas the Kaiko family leads to the lowest performance. However, since the mean AUROC5 performance of the analyzed VFMs converges and lies within 3 pp 0·71 to 0·74 when the full training dataset is utilized, the number of training samples seems to be more important than the actual VFM that is used.

**Figure 3:**
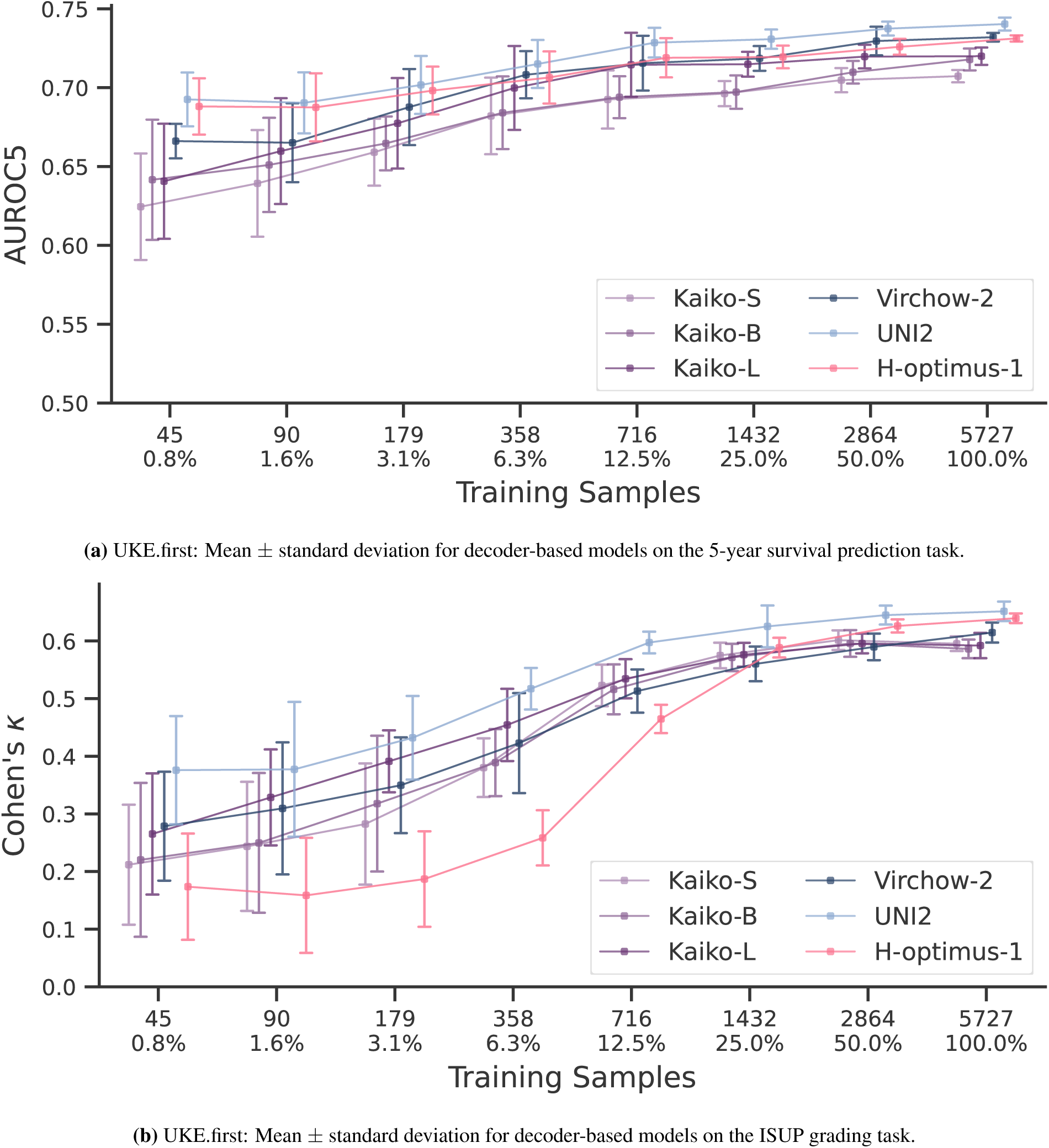
Saturation analysis: Mean ± standard deviation for the foundation models on 5-year relapse prediction (a) and ISUP classification (b). The x-axis displays the number of training samples on a logarithmic scale, computed from five random sub-samples of training samples.

We observe similar findings for the ISUP classification task (see fig. 3b and table B.6b). More training data leads to higher performance and lower standard deviation, a trend that applies to all analysed foundation models. The Kaiko models demonstrate comparable performance for only 50 % of training data (*n* = 2864) compared to all 5727 samples, with Cohen’s *κ* values ranging from 0·59 to 0·6 indicating model saturation. The larger models achieve the best performance ranging from 0·62 for Virchow-2 to 0·65 for UNI2. The results on the full training set compared to only *n* = 45 training samples show that Cohen’s *κ* has improved for all models by 27 to 47 pp. While Virchow-2 and UNI2 benefited less from increases in training dataset size, a significant amount of 1432 training samples was necessary for H-optimus-1 to achieve better Cohen’s *κ* compared to the three smaller Kaiko models. For more than 1432 training samples, the performance of H-optimus-1 becomes comparable to that of UNI2, improving its performance by 47 pp. Similarly to 5-year relapse prediction, UNI2 achieves the highest overall results for all training data subsets. Further, the standard deviation decreases with larger datasets for all models. With a deviation ranging from 0·092 to 0·134 for 45 training samples, it reaches values between 0·008 to 0·022 for 5727 training samples.

These findings suggest that the analysed VFMs require large datasets with ∼1000 training samples for 5-year relapse prediction and ∼2000 training samples for ISUP classification with high fidelity. While larger models seem to outperform smaller ones when little training data is available, data requirements for peak performance are similar.

### 3.3. VFMs are negatively affected by domain shift

This section evaluates the performance of the different VFMs on previously unseen variations in the digitization protocol, as detailed in section 2.2. Each model was tested on TMAs representing distinct acquisition conditions while maintaining consistent patient-level information. In total, up to six TMAs per patient were used, capturing differences in scanner type, staining time and section thickness. Ideally, model predictions should remain stable under such variations. However, notable performance degradation was observed across all VFMs as illustrated in fig. 4 and table B.7.

**Figure 4:**
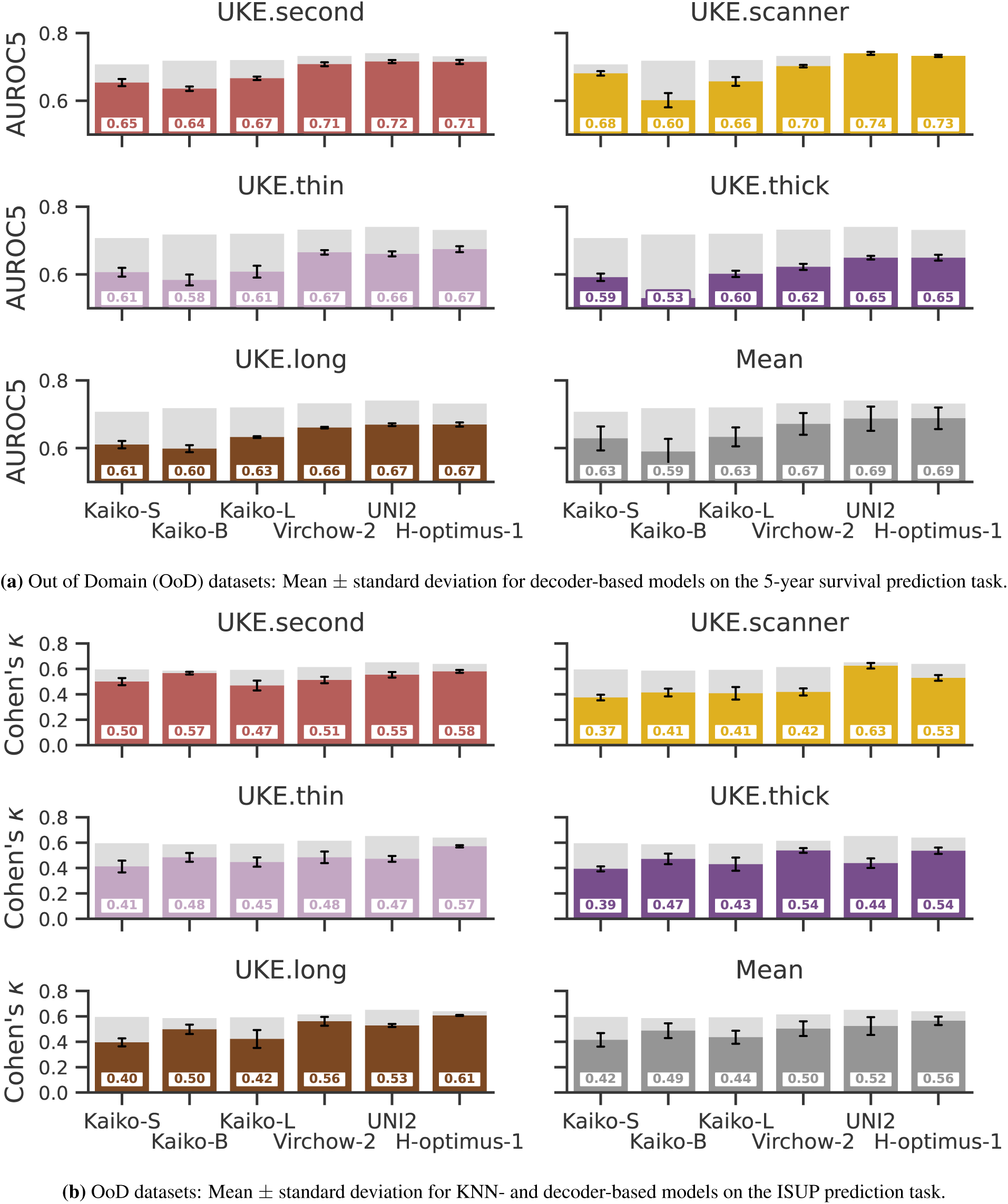
Robustness Analysis: Comparing mean ± standard deviation of baseline (UKE.first, light gray) performance of VFMs on the five other, potential OoD datasets for 5-year relapse prediction (a) and ISUP classification (b). The last plot represents the mean OoD results, that shows the mean performance for one model over all OoD datasets.

We evaluate the VFMs on the five different variations in the sample preparation protocol as described in section 2.2 and compare the results to their initial performance on our main sub-dataset UKE.first that we now use as a baseline. The VFMs showed strongest performance on this sub-dataset in 5-year relapse prediction (AUROC5 between 0·71 for Kaiko-S and 0·74 for UNI2) and ISUP classification (Cohen’s *κ* between 0·59 for Kaiko-S and Kaiko-B and 0·65 for UNI2). Notably, UNI2 and H-optimus-1 showed similar AUROC5 performance (0·74 for UNI2 and 0·73 for H-optimus-1) on the UKE.scanner sub-dataset.

Evaluating the six VFMs on the five sub-datasets on 5-year relapse prediction in terms of AUROC5 reveals a general trend where the three larger models, namely Virchow-2, UNI2, and H-optimus-1 show a smaller performance drop compared to the three smaller Kaiko models. Exemplary on UKE.second, the larger models show a slightly lower performance of 2pp, while Kaiko-S, Kaiko-B, and Kaiko-L lose 6 pp, 8 pp, and 5 pp respectively. Notably, all models perform worst on UKE.thick. While the smaller models lose 11 pp, 19 pp, and 12 pp for Kaiko-S, Kaiko-B, Kaiko-L respectively, Virchow-2, UNI2, and H-optimus-1 experience a loss of 11 pp, 9 pp, and 8 pp respectively. Comparing the mean performance among all sub-datasets except UKE.first, Kaiko-B performs worst with an average AUROC5 of 0·59 while both UNI2 and H-optimus-1 score an average AUROC5 of 0·69 which is still 3 pp to 4 pp below their scores for UKE.first.

For ISUP classification, a similar trend can be observed (see fig. 4b and table B.7b). All VFMs perform best on the UKE.first sub-dataset that was used for training, with a Cohen’s *κ* of 0·59 to 0·60 for the three Kaiko models and 0·62, 0·65, and 0·64 for Virchow-2, UNI2, and H-optimus-1 respectively. Further, all models experience a significant drop in performance for all analysed sub-datasets with variations in the digitization protocol. While Kaiko-S, Kaiko-B, and Kaiko-L drop by 18 pp, 10 pp, and 16 pp respectively in their mean performance compared to UKE.first, Virchow-2, UNI2, and H-optimus-1 experience a drop of 12 pp, 13 pp, and 7 pp respectively. In contrast to the 5-year relapse evaluation of UKE.scanner where UNI2 and H-optimus-1 maintained baseline performance, it posed significant problems for all VFMs in ISUP classification with a drop of 11 pp to 21 pp for all models with the notable exception of UNI2 that only showed a drop of 2 pp. Except for UKE.scanner, H-optimus-1 is superior to UNI2. The mean loss in performance in terms of Cohen’s *κ* is significant among all models ranging from 12 pp to 18 pp for all models with the exception of H-optimus-1 that only shows a drop of 7 pp.

To summarize, the results for 5-year relapse prediction and ISUP classification demonstrate a significant decrease in performance for all sub-datasets demonstrating a lack of robustness in all analysed VFMs. Larger foundation models are generally more robust to domain shift, which is particularly true for the easier task of relapse prediction.

## 4. Discussion

In this study, we conducted a systematic evaluation of six contemporary VFMs under controlled yet realistic protocol variants. Using a large, protocol-variant prostate cancer TMA dataset, we examined the impact of tissue sampling location, scanner vendor, variations in section thickness, and staining duration. These are dimensions of variability that reflect everyday practice but are rarely analyzed beyond simple scanner changes as provided in.^8^ To the best of our knowledge, only our previous studies have examined controlled protocol-level variation at similar depth.^10, 11^

Our results yield three central observations. First, while VFMs provide rich representational features, downstream performance depends strongly on the chosen decoder architecture (section 3.1). Simple probing approaches such as KNN, which are commonly used in the evaluation of foundation models,^1, 14, 22^ were insufficient for clinically relevant tasks. Second, the presumed data efficiency of VFMs did not hold: stable decoder performance typically required more than 1000 training samples (see section 3.2). Third, none of the models demonstrated sufficient robust generalization under protocol-level domain shifts as shown in section 3.3. Even with comparatively large training sets, performance decreased substantially across all shift types (ISUP classification with a 10 pp to 20 pp drop in terms of Cohen’s *κ*, 5-year relapse prediction with 4 pp to 13 pp in AUROC5), underscoring that current VFM pipelines remain sensitive to routine protocol variation.

These findings challenge assumptions regarding inherent VFM robustness, especially for the larger analysed models.^1, 2, 16^ This work highlights a persistent gap between benchmark performance and the reliability required for clinical deployment. Notably, scaling model size alone did not yield emergent robustness: even the largest models tested failed to generalize under protocol variation. Increasing the training dataset to 5727 TMAs saturated 5-year relapse prediction performance but did not improve robustness under protocol shift. The observed performance degradation reflects a broader limitation of current VFMs: the inability to reliably disentangle biological morphology from protocol-dependent stylistic variation. Similar challenges have been reported in studies that quantify domain influence in the embedding space,^6, 23^ suggesting structural fragility in current representation learning approaches.

Addressing these limitations will require advances at multiple levels. From a training perspective, including model robustness directly, for instance domain-adversarial learning,^24, 25^ or targeted augmentation strategies^26, 27^ may help build more domain-invariant representations. Stain-aware pretraining, multi-domain or multi-institutional self-supervised learning, and augmentations designed to mimic real-world technical variability may further enhance representation stability. We also deliberately froze the VFM encoder networks to isolate representational robustness. While this choice prevents feature extractor overfitting to the training domains and downstream-specific tasks, it may underestimate the potential benefits of end-to-end fine-tuning.^28^ Decoder design plays a central role in how effectively VFM embeddings are utilized. More expressive decoder architectures, such as cross-attention,^29, 10, 11^ or MIL approaches beyond ABMIL^20^ could better leverage VFM embeddings and improve robustness under domain shift.^30, 31, 32^ Building on this, relaxing the frozen-encoder assumption through layer unfreezing, low-rank adaptation,^28^ or other parameter-efficient tuning methods^33^ may enable more flexible domain adaptation with reduced risk of catastrophic forgetting. Additionally, newer self-supervised backbones such as `DINOv3`^22^ may provide more domain-invariant features than currently used models.

While we assume that our results provide general insights into the data requirements, robustness, and performance of state-of-the-art VFMs for histopathology, we also acknowledge potential shortcomings of our work. We believe that task selection, for instance, could affect the conclusions made in this manuscript. We included one comparatively complex task (ISUP classification) and one simplified prognostic task (5-year relapse prediction). Wider task variation in future work could further clarify how task complexity shapes VFM robustness and data requirements. Furthermore, TMAs represent standardized and spatially constrained tissue regions that differ substantially from full WSIs that are standard for clinical diagnosis. Robustness patterns observed in TMAs may underestimate the additional challenges that are posed by whole-slide heterogeneity. Lastly, this study relies on a single institution prostate cancer dataset, which might not capture the variability induced by other centres’ staining practices, tissue handling, or scanner calibration routines. Broader and more diverse datasets are an essential next step. Multi-institution cohorts spanning heterogeneous digitization protocols, scanner vendors, and tissue-processing standards will be critical for VFMs to generalize reliably across clinical settings.^34^

## Computational hardware and software

The project was implemented using python 3.11.11 with pytorch-lightning 1.9.5 The decoders for the VFMs were trained on a NVIDIA Quadro RTX 8000 with 48GB GPU memory and an Intel(R) Xeon(R) Silver 4214 CPU.

## Contributors

SB and GS initialized this work. SB, MZ, GS, PF, and AW conceptualized the project, algorithm, and computational analyses. SB, MZ, PF, EB, TBH, JB, and GS supervised the work. GS and ML collected and aggregated the data. AW and PF were responsible for the computational data analysis and algorithmic adaptations. AW, PF, MZ, and SB wrote the manuscript. All authors critically read and amended the manuscript. Funding was acquired by SB, MZ, JB, and TBH.

## Disclosure

During the preparation of this work the author(s) used the GPT4.1 model of UHHGPT (https://uhhgpt.uni-hamburg.de/), in order to improve the creative writing process. After using this service, the authors reviewed and edited the content as needed and take full responsibility for the content of the published article.

## Data

The use of archived remnants of diagnostic tissues for manufacturing of TMAs and their analysis for research purposes as well as patient data analysis has been approved by local laws (HmbKHG, paragraph 12) and by the Ethics commission Hamburg (WF-049/09). All work has been carried out in compliance with the Helsinki Declaration. The dataset can be obtained upon reasonable request to the department of Pathology at the UKE.

## Declaration of interests

PF, SB and EB work part time for Spearpoint Analytics AB, a company developing AI-based digital pathology solutions. The authors declare no other conflicts of interest.

## Data Availability

The data is available upon reasonable request from the Department of Pathology of the University Medical Center Hamburg-Eppendorf.

## Acknowledgments

We would like to thank the IT of the Institute for Medical Systems Bioinformatics and the bAIome Center for Biomedical AI, Sven Heins and Vadim Ustinov.

PF was supported by DFG SFB 1286 SP02. SB received funding from DFG SFB 1713 (C01), SFB 1700 (SP01), and TRR 422 (CP2). JB received funding from DFG SFB 1700 SP01. TBH was supported by DFG SFB 1192 B8. MZ received funding from DFG SFB 1192 B9. MZ and AW received funding from DFG SFB 1700 A8. AW, JB, and SB were supported by CDL FLIGHT of the University of Hamburg.

## Appendix A. Methods

Appendix A.1. Data

**Table A.2:** Patient metadata across the analyzed sub-datasets.

|  |  | UKE.first | UKE.second | UKE.scanner | UKE.thin | UKE.thick | UKE.long |
| --- | --- | --- | --- | --- | --- | --- | --- |
|  | Count | 1208 | 1164 | 1208 | 885 | 891 | 893 |
| Age | (median) | 64.4 | 64.3 | 64.4 | 63.9 | 64.0 | 64.1 |
| PSA Level | IQR | [4.70, 11.42] | [4.76, 11.57] | [4.70, 11.42] | [4.65, 10.88] | [4.69, 10.89] | [4.69, 11.00] |
| Censored |  | 57.8% | 57.9% | 57.8% | 66.3% | 66.4% | 66.9% |
| Median Survival | (months) | 23.8 | 23.5 | 23.8 | 30.2 | 30.5 | 30.2 |
| Median Followup | (months) | 96.0 | 96.0 | 96.0 | 91.0 | 91.0 | 91.0 |
| ISUP | 0 | 71 | 69 | 71 | 63 | 63 | 64 |
|  | 1 | 224 | 219 | 224 | 171 | 180 | 177 |
|  | 2 | 610 | 584 | 610 | 473 | 471 | 471 |
|  | 3 | 231 | 224 | 231 | 141 | 140 | 146 |
|  | 4 | 24 | 21 | 24 | 16 | 15 | 14 |
|  | 5 | 48 | 47 | 48 | 21 | 22 | 21 |
| Event Type | FU | 698 | 674 | 698 | 587 | 592 | 597 |
|  | BCR | 506 | 486 | 506 | 297 | 298 | 295 |
|  | META | 4 | 4 | 4 | 1 | 1 | 1 |
| path. T-Stage | ≤ T2 | 691 | 666 | 691 | 544 | 540 | 541 |
|  | T3 | 501 | 482 | 501 | 332 | 340 | 341 |
|  | T4 | 16 | 16 | 16 | 9 | 11 | 11 |

**Table A.3:**
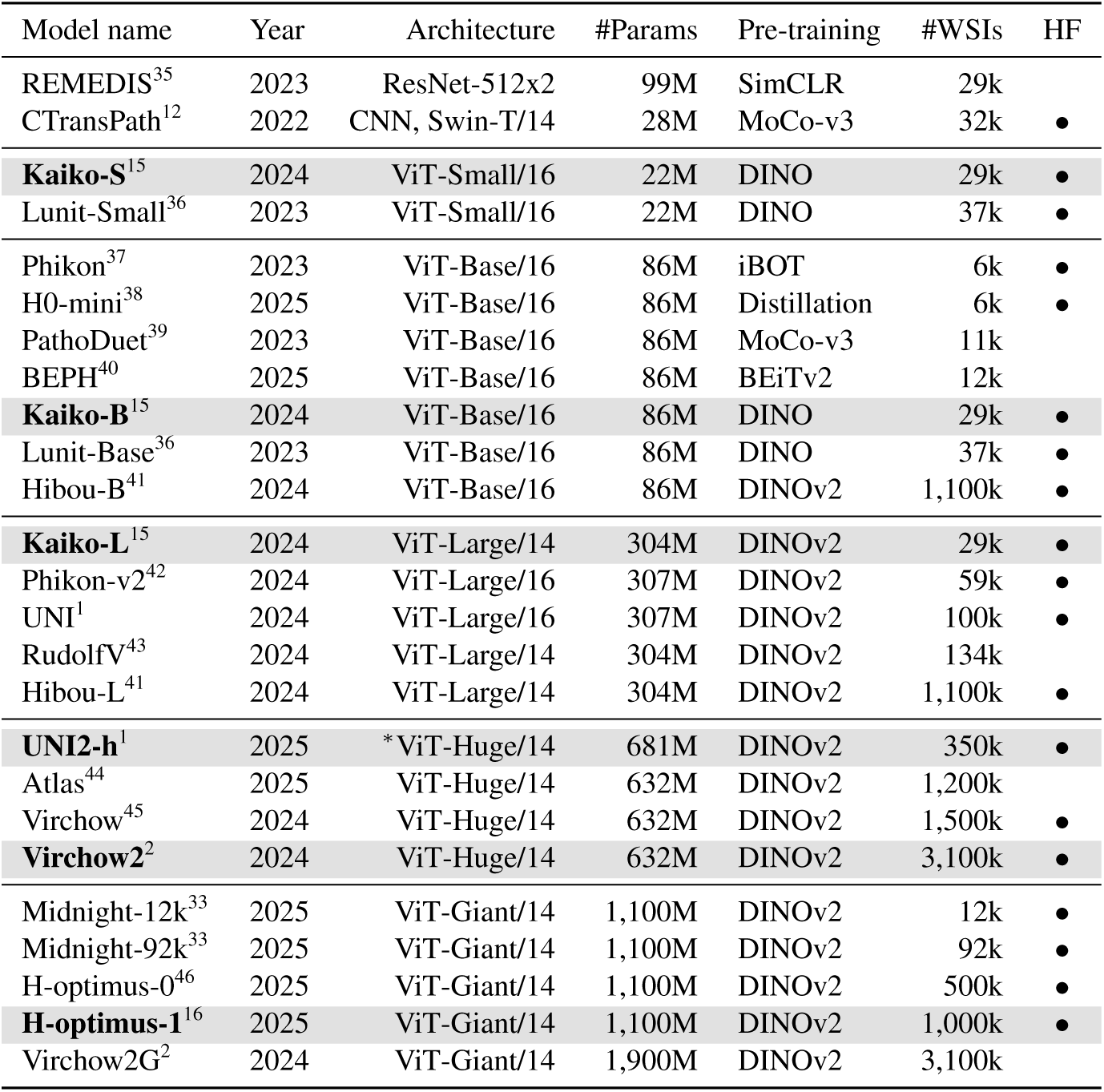
Overview of vision foundation models for computational pathology, that is sorted by *Architecture* and *#WSIs*. Bold model names with shaded rows were selected for our experiments. Architectures with ^∗^ have modified numbers of layers or attention heads. *#Params* represents the number of parameters for each VFM, that has been pre-trained with a specific number of pre-training images (*#WSIs*). *HF* refers to the availability of model weights on https://huggingface.co/.

**Table A.4:**
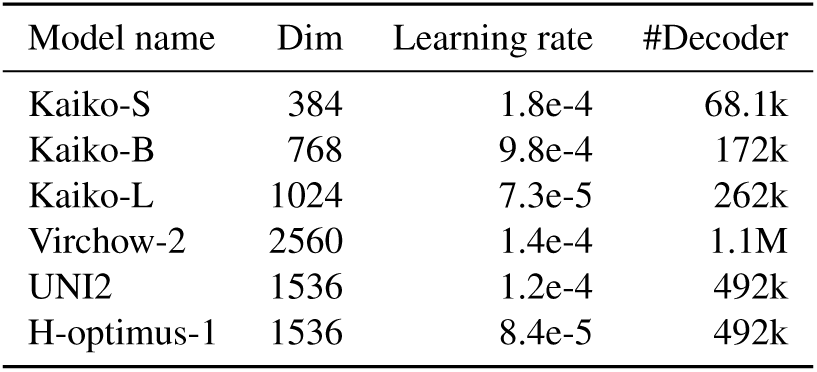
Training parameters of the compared vision foundation models. *Dim* specifies the output embedding dimension for each VFM and *#Decoder* shows the number of trainable decoder parameters for each architecture.

## Appendix B. Results

Appendix B.1. KNN vs. Decoder

**Table B.5:** KNN vs decoder results (mean ± standard deviation) for all models on UKE.first. The best value per model is highlighted in bold.

| Method | Kaiko-S | Kaiko-B | Kaiko-L | Virchow-2 | UNI2 | H-optimus-1 |
| --- | --- | --- | --- | --- | --- | --- |
| KNN | 0.557 ± 0.002 | 0.564 ± 0.013 | 0.569 ± 0.007 | 0.583 ± 0.012 | 0.584 ± 0.009 | 0.586 ± 0.005 |
| Decoder | **0.699 ± 0.009** | **0.711 ± 0.010** | **0.716 ± 0.005** | **0.733 ± 0.007** | **0.737 ± 0.004** | **0.730 ± 0.003** |
| (a) Area under the Receiver Operating Characteristic Curve for 5-year relapse prediction |  |  |  |  |  |  |
| Method | Kaiko-S | Kaiko-B | Kaiko-L | Virchow-2 | UNI2 | H-optimus-1 |
| KNN | 0.104 ± 0.021 | 0.129 ± 0.031 | 0.109 ± 0.029 | 0.184 ± 0.036 | 0.216 ± 0.035 | 0.194 ± 0.034 |
| Decoder | **0.593 ± 0.015** | **0.571 ± 0.031** | **0.590 ± 0.021** | **0.588 ± 0.014** | **0.640 ± 0.014** | **0.628 ± 0.011** |
| (b) Cohen's $\kappa$ for ISUP prediction | | | | | | |

Appendix B.2. Saturation Analysis

**Table B.6:** Saturation Analysis (mean ± standard deviation) for all models on UKE.first. The best value per model is highlighted in bold, the second best with an underscore.

| #Samples | Model Ratio | Kaiko-S | Kaiko-B | Kaiko-L | Virchow-2 | UNI2 | H-optimus-1 |
| --- | --- | --- | --- | --- | --- | --- | --- |
| 45 | 0.8% | 0.625 ± 0.034 | 0.642 ± 0.038 | 0.641 ± 0.036 | 0.666 ± 0.011 | 0.693 ± 0.017 | 0.688 ± 0.018 |
| 90 | 1.6% | 0.639 ± 0.034 | 0.651 ± 0.030 | 0.660 ± 0.034 | 0.665 ± 0.025 | 0.690 ± 0.019 | 0.688 ± 0.022 |
| 179 | 3.1% | 0.659 ± 0.021 | 0.665 ± 0.017 | 0.677 ± 0.029 | 0.688 ± 0.024 | 0.702 ± 0.018 | 0.698 ± 0.015 |
| 358 | 6.3% | 0.682 ± 0.024 | 0.684 ± 0.023 | 0.700 ± 0.027 | 0.708 ± 0.015 | 0.715 ± 0.015 | 0.706 ± 0.016 |
| 716 | 12.5% | 0.693 ± 0.018 | 0.694 ± 0.013 | 0.715 ± 0.020 | 0.716 ± 0.017 | 0.729 ± 0.009 | 0.719 ± 0.012 |
| 1432 | 25.0% | 0.696 ± 0.008 | 0.697 ± 0.011 | 0.715 ± 0.008 | 0.719 ± 0.008 | 0.731 ± 0.006 | 0.719 ± 0.007 |
| 2864 | 50.0% | 0.705 ± 0.008 | 0.710 ± 0.007 | **0.720 ± 0.007** | 0.730 ± 0.009 | 0.737 ± 0.004 | 0.726 ± 0.005 |
| 5727 | 100.0% | **0.707 ± 0.004** | **0.718 ± 0.007** | 0.720 ± 0.005 | **0.732 ± 0.003** | **0.740 ± 0.004** | **0.731 ± 0.002** |
| (a) Area under the Receiver Operating Characteristic Curve for 5-year relapse prediction |  |  |  |  |  |  |  |
| #Samples | Model Ratio | Kaiko-S | Kaiko-B | Kaiko-L | Virchow-2 | UNI2 | H-optimus-1 |
| 45 | 0.8% | 0.212 ± 0.104 | 0.220 ± 0.134 | 0.265 ± 0.105 | 0.279 ± 0.094 | 0.376 ± 0.094 | 0.174 ± 0.092 |
| 90 | 1.6% | 0.244 ± 0.112 | 0.250 ± 0.121 | 0.329 ± 0.083 | 0.310 ± 0.115 | 0.377 ± 0.117 | 0.159 ± 0.100 |
| 179 | 3.1% | 0.282 ± 0.105 | 0.318 ± 0.118 | 0.391 ± 0.054 | 0.350 ± 0.083 | 0.432 ± 0.073 | 0.187 ± 0.083 |
| 358 | 6.3% | 0.380 ± 0.051 | 0.389 ± 0.058 | 0.454 ± 0.063 | 0.423 ± 0.087 | 0.517 ± 0.036 | 0.258 ± 0.048 |
| 716 | 12.5% | 0.523 ± 0.036 | 0.516 ± 0.043 | 0.534 ± 0.034 | 0.513 ± 0.038 | 0.597 ± 0.019 | 0.465 ± 0.025 |
| 1432 | 25.0% | 0.575 ± 0.022 | 0.571 ± 0.024 | 0.576 ± 0.021 | 0.560 ± 0.030 | 0.625 ± 0.036 | 0.588 ± 0.017 |
| 2864 | 50.0% | **0.601 ± 0.017** | **0.596 ± 0.023** | **0.596 ± 0.017** | 0.590 ± 0.023 | 0.645 ± 0.016 | 0.626 ± 0.011 |
| 5727 | 100.0% | 0.595 ± 0.013 | 0.586 ± 0.016 | 0.592 ± 0.022 | **0.615 ± 0.017** | **0.651 ± 0.017** | **0.639 ± 0.008** |
| (b) Cohen's $\kappa$ for ISUP prediction | | | | | | | |

Appendix B.3. Robustness Analysis

**Table B.7:**
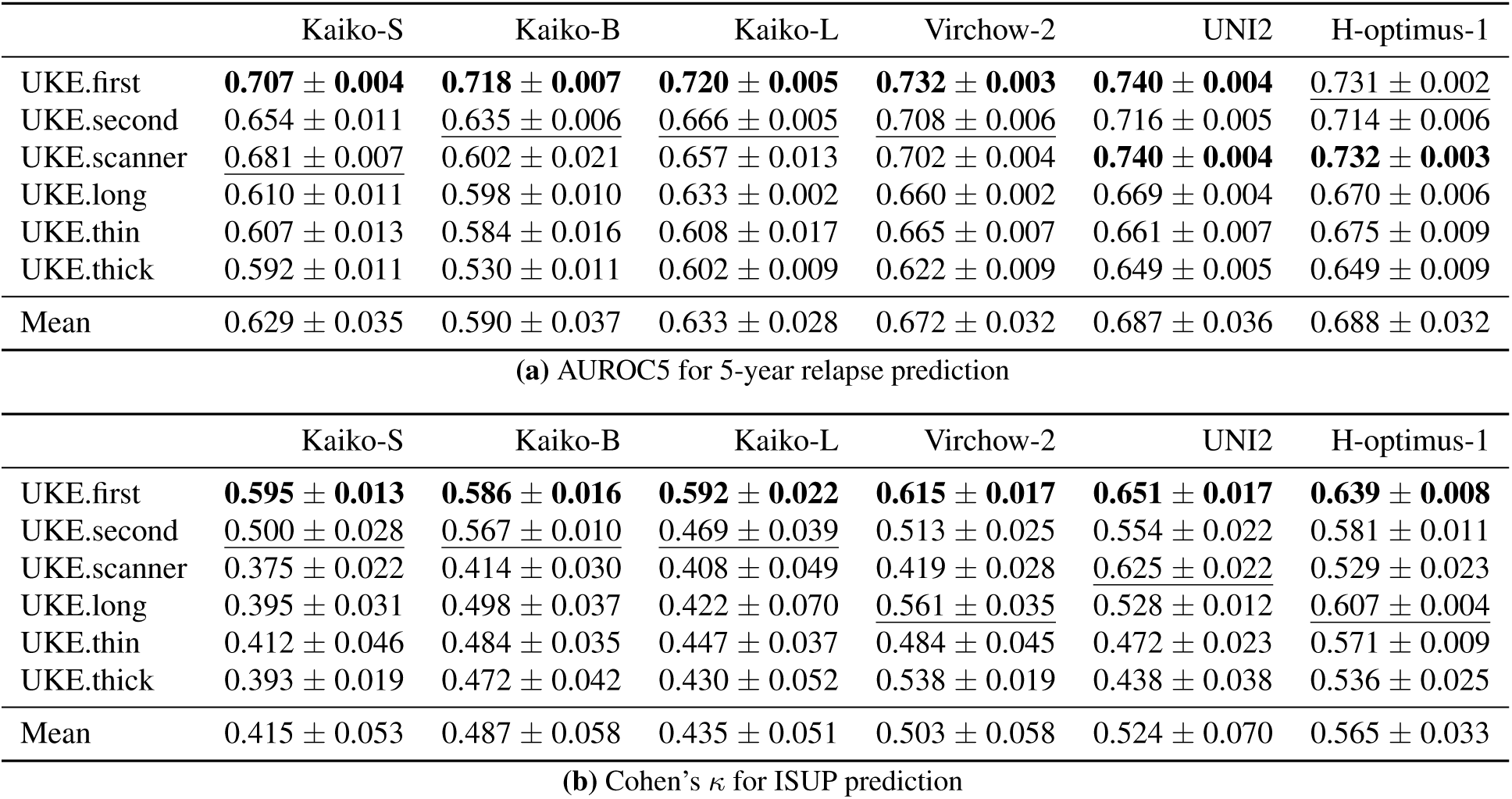
Robustness evaluation results (mean ± standard deviation) for all models and datasets. The best value per model is highlighted in bold, the second best with an underscore. ŌōD̄ = Mean performance on OoD datasets.

## Footnotes

3 https://huggingface.co/

## Notes

### Author Declarations

The use of archived remnants of diagnostic tissues for manufacturing of TMAs and their analysis for research purposes as well as patient data analysis has been approved by local laws (HmbKHG, paragraph 12) and by the Ethics commission Hamburg (WF-049/09). All work has been carried out in compliance with the Helsinki Declaration.

